# The relationship between limb dystonia severity and functional impact in children with cerebral palsy

**DOI:** 10.64898/2026.04.11.26350684

**Authors:** Emma Lott, Susie Kim, Joanna Blackburn, Rose Gelineau-Morel, Dararat Mingbunjerdsuk, Jennifer O’Malley, Laura Tochen, Jeff L. Waugh, Steve Wu, Bhooma R. Aravamuthan

## Abstract

Dystonia treatment evaluation in cerebral palsy (CP) is limited by the lack of clinician-assessed scales linking dystonia severity to functional impact. We asked 7 pediatric movement disorder specialists to review videos of 27 children with CP while performing an upper extremity task and while walking. Experts rated arm and leg dystonia severity using the Global Dystonia Severity Rating Scale (GDRS) and task-specific functional impact on a five-point scale adapted from the Dyskinetic Cerebral Palsy Functional Impact Scale. Arm GDRS scores correlated with functional impact on the upper extremity task (linear regression R^2^=0.48, p=0.0005). Leg GDRS scores correlated with gait impact (R^2^=0.43, p=0.001). A four-point increase in total GDRS corresponded to a one-point worsening in combined functional impact. By demonstrating how expert-rated limb dystonia severity correlates with task-specific functional impact in children with CP, these results could help clinically identify functionally-meaningful differences in dystonia severity.

## Introduction

Dystonia is a debilitating movement disorder characterized by involuntary movements triggered by attempted voluntary movement, handling, or stress.^1–3^ In children, dystonia is typically seen in association with cerebral palsy (CP), the most common childhood-onset motor disability worldwide.^4–7^ Dystonia in CP is common, affecting at least 1 of every 1000 people.^4,6,7^ However, there is little evidence to support any treatments for dystonia in CP.^8^ Treatment evaluation is significantly limited by the lack of functionally meaningful dystonia assessment scales.^9^ The caregiver-assessed Dyskinetic Cerebral Palsy Functional Impact Scale (D-FIS) was recently developed but requires significant caregiver knowledge to both identify dystonia in their child across multiple tasks and differentiate dystonia from many other co-existing motor symptoms.^10^ This pre-requisite caregiver knowledge significantly limits the practical clinical use of the D-FIS. Clinician-assessed scales that rate dystonia severity and are validated in people with CP assess factors evident on visual assessment (e.g. the amplitude and duration of dystonic movements) but may or may not additionally consider the functional impact of these movements.^11–13^ Furthermore, existing clinician-assessed scales use the summation of dystonia severity scores across all body regions as a single score, which dilutes dystonia severity scores for a single body region that may still be functionally impactful, even if the whole-body dystonia severity score remains low.^14^ Overall, it remains unclear how or whether clinician-assessed dystonia severity scales correlate with clinician-assessed functional impact on motor tasks.

Minimum clinically important differences for these scales have not been calculated in any context, to the best of our knowledge, and have not been published with expert clinician-assessed functional impact as a diagnostic gold standard. Responsiveness has been calculated for dystonia severity scales but only in the minority of children with CP who have dystonia as their predominant type of motor tone abnormality and/or with dystonia severe enough to warrant surgical interventions.^11,15,16^ Therefore, the ability of these scales to detect functionally meaningful differences in children with CP, most of whom have milder dystonia severity, remains unknown.^6,17^ This unassessed majority of children with CP and dystonia may not be eligible for surgical interventions but may still have functionally limiting dystonia that warrants pharmacologic or injectable interventions like botulinum toxin.^8,18^ Therefore, we must establish whether clinician-assessed dystonia severity scales can detect functionally meaningful differences in dystonia severity for this majority of children with CP.

To address this knowledge gap, we asked pediatric movement disorders physicians to use the Global Dystonia Severity Rating Scale (GDRS)^19^ to rate dystonia severity in the arms and legs of children with CP using videos of these children while performing an upper extremity task and while walking. We also asked these physicians to rate how dystonia functionally impacted the performance of each task. We hypothesized that dystonia severity ratings in the arms vs. the legs would differently correlate with functional impact in the upper extremity task vs. walking. We also used this data to determine what score difference on the GDRS correlated with a clinician-assessed difference in dystonia’s functional impact on a task, thus calculating an MCID for the GDRS using clinician-assessed functional impact as the anchor. Our goal is to use these results to facilitate the functionally meaningful use of dystonia severity rating scales like the GDRS in clinical treatment trials.

## Methods

We received Washington University School of Medicine Institutional Review Board approval for this project (Approval Number: 202207105; Date of Approval 12/02/2024).

### Subjects and tasks

Subjects were recruited from the pool of children cared for at a large tertiary care CP center (St. Louis Children’s Hospital, St. Louis, MO) between 9/21/23 and 1/18/24. Inclusion criteria were a clinician-confirmed diagnosis of CP, ages 4 to <18 years old, and the ability to independently attempt a seated alternating hand open-close task and a walking task as identified during videos of these tasks taken during routine clinical care. Age 4 was used as the lower limit for inclusion to ensure that children had the cognitive ability and attention span to complete the tasks.

For the hand open-close task, the child was seated on a tall chair with their legs dangling and their hands resting on their lap. The child was asked to raise their dominant hand and alternately open and close (fist) that hand as quickly as possible for 5 seconds. Afterward, the child placed their dominant hand back on their lap and raised their non-dominant hand, repeating the open-close motion. This process was repeated three times. For the walking task, the child initially started by standing and facing the camera. They then turned away from the camera and walked 10 feet, then turned around and walked 10 feet back towards the camera. For both tasks, the child had their shirt sleeves and pants rolled up to show the elbows and knees. Videos were recorded using a tripod-mounted Google Pixel 3a smartphone at 1920×1080 pixel resolution at 30 frames/second.

Demographics for all subjects (age, sex, gestational age at birth, tone/movement disorder types, CP distribution, Gross Motor Function Classification System Level,^20^ and Manual Ability Classification System Level^21^) were automatically extracted from clinical documentation as we have previously described.^6^ Notably, in-clinic determinations regarding the presence or absence of dystonia were done by the treating CP center clinician (either a fellowship-trained pediatric movement disorders physician or a dedicated subspeciality nurse practitioner) using a standardized assessment incorporating the Hypertonia Assessment Tool and caregiver-based dystonia screening.^22^

### Expert video review

To ensure subject anonymity, faces of the subjects and any caregivers (if present) were blurred using open-source software (ShotCut, Meltytech, LLC) and custom written software (SecurePose).^23^

Experts are all fellowship-trained pediatric movement disorders physicians (RGM, DM, JOM, JW, SW, JB, LT) from seven different clinical sites across the country with established expertise in determining dystonia severity ratings from consensus-based video review of children with CP.^24,25^ Experts were aware that all subjects had been diagnosed with CP but were given no other information about the subjects. Experts first independently reviewed anonymized and face-blurred videos of children with CP doing the alternating hand open-close task and the walking task. Experts then met virtually to review and discuss the videos of all subjects (Zoom Video Communications) and arrive at consensus-based agreement for the dystonia severity ratings (GDRS) and functional impact ratings (adapted from the D-FIS). ^10,19^ After this discussion, experts entered their scores via REDCap forms and the median score across all 7 experts was used for further analysis.

The GDRS was chosen because it is the least prescriptive of dystonia severity rating scales used to assess children with CP and thus allows the raters to globally consider features that impact dystonia severity. We have shown that experts consider amplitude, duration/frequency, and functional impact when providing GDRS ratings for children with CP.^26^ Therefore, unlike other more prescriptive dystonia severity rating scales, the GDRS may be particularly well-suited to estimate the functional impact of dystonia in a particular body region.^24–26^ Experts used the GDRS only to rate arm and leg dystonia because the motor tasks assessed relied primarily on limb function. Additionally, face-blurring to ensure anonymity prevented accurate head/neck/trunk dystonia severity assessment (with experts noting that the occasional blurring of the upper half of the neck made it difficult to accurately gauge trunk movement). The GDRS asks experts to rate each limb segment (proximal and distal) on at 10-point Likert scale (0 – no dystonia, 1 – minimal dystonia, 5 – moderate dystonia, 10 – most severe dystonia). This yields a possible GDRS score range of 0-40 for the arms (proximal and distal left and right limb) and 0-40 for the legs.^19^

Experts rated dystonia’s functional impact on each task using a 5-point Likert scale (Table 1) adapted from the caregiver-assessed D-FIS.^10^ The D-FIS was used because no existing clinician-assessed scale is specifically dedicated to the assessment of the functional impact of dystonia.

**Table 1.** Wording for dystonia functional impact assessment for the hand open-close and gait tasks, as adapted from the caregiver-facing Dyskinetic Cerebral Palsy Functional Impact Scale.^10^

| Score | Description |
| --- | --- |
| 0 | Dystonia DOES NOT affect this task. |
| 1 | Dystonia makes it a LITTLE HARDER to do this task. |
| 2 | Dystonia makes it a LOT HARDER to do this task; Dystonia makes it harder to do this task on their own or makes it harder for others to help this child do this task. |
| 3 | Dystonia makes it EXTREMELY HARD to do this task; Dystonia makes this child need support from another person to do this task, and caregivers must have experience with this movement to help this child do this task. |
| 4 | Dystonia makes it IMPOSSIBLE to do this task even with help. |

### Statistical analysis

All statistical analyses were done in GraphPad Prism (version 10, GraphPad Software LLC). Chi-square tests and t-tests were used to compare demographics between eligible subjects who met inclusion criteria and the subjects who ultimately participated in the study. T-tests were used to determine any differences in experts’ GDRS or functional impact ratings between subjects who did or did not have dystonia based on their routine in-clinic evaluations. Linear regression was used to determine correlations between experts’ consensus scores between the arm GDRS subscore, leg GDRS subscore, hand open-close functional impact rating, and gait functional impact rating, with Bonferroni corrections for multiple comparisons. The GDRS score difference corresponding to a change in clinician-assessed functional impact was calculated as the slope of the linear regression line determining the correlation between the summed arm and leg GDRS scores and the summed hand open-close and gait functional impact ratings.^27^ The significance level was set *a priori* at p<0.05.

## Results

During the recruitment period, 77 children met inclusion criteria. Of these 77 eligible children, the caregivers/guardians of 27 (35%) consented to participate in this study. The demographics of children eligible for the study (n=77) and those who participated (n=27) are shown in Table 2. There was no significant difference in these demographics between those eligible and those who participated (p>0.05, Chi-square tests and t-tests, Table 2). Most participating subjects were able to independently ambulate without assistive devices (21/27, 78%, at Gross Motor Function Classification System Levels I-II) and were able to hold and use objects independently (25/27, 93%, at Manual Ability Classification System Level I-II).

**Table 2.**
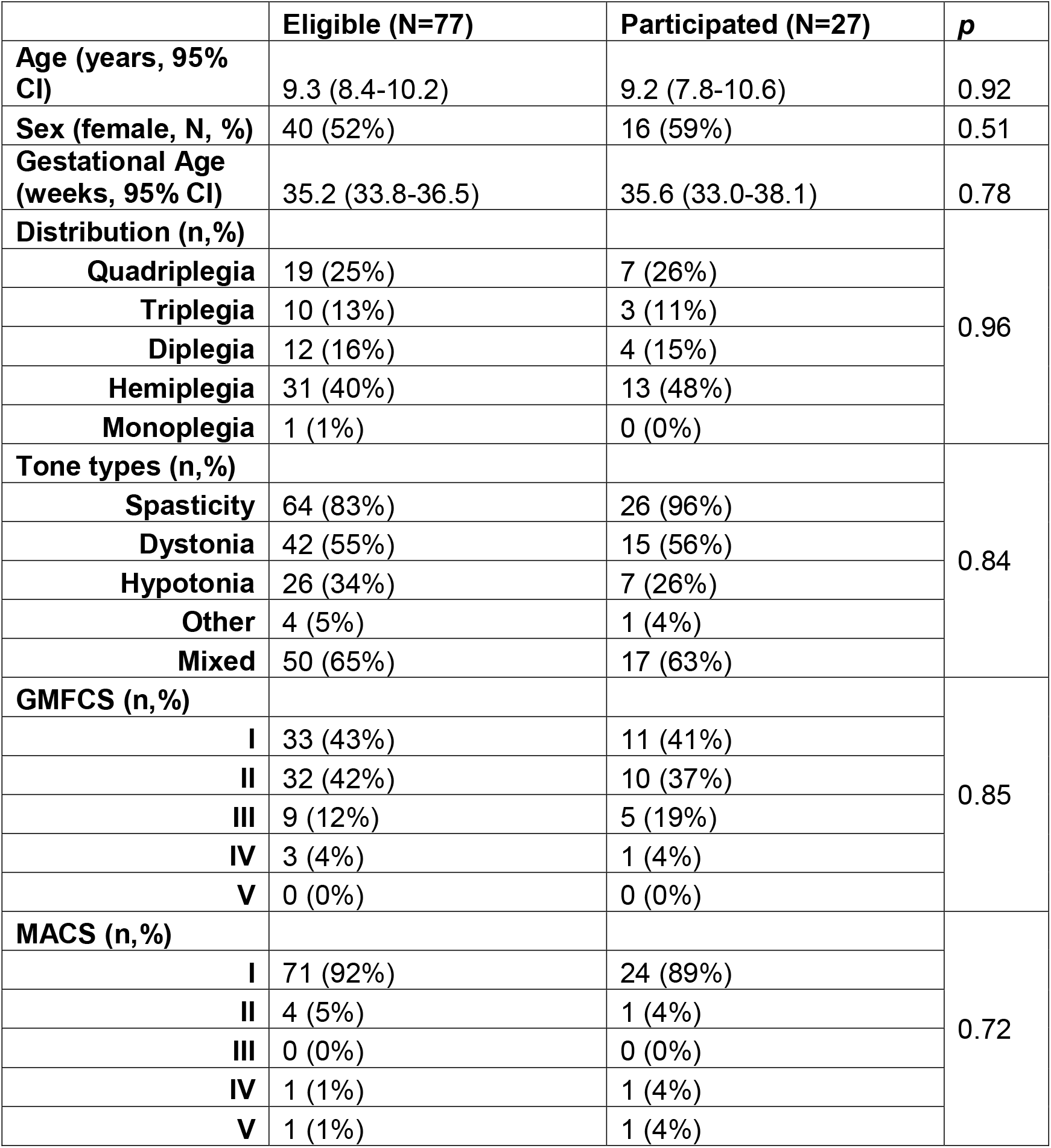
Subject demographics of those eligible and those participating in the study. GMFCS: Gross Motor Function Classification System (Level I-II: independently ambulatory without assistive devices, Level III: independently ambulatory with an assistive device, Level IV-V: primarily uses a wheelchair),^20^ MACS: Manual Ability Classification System (Level I-II: holds and uses objects independently, Level III: uses objects with help from others, Level IV: uses some objects with help from others, Level V: does not hold/use objects).^21^ Other tone type includes athetosis, chorea, and ataxia. Mixed tone type accounts for subjects with more than one of spasticity, dystonia, hypotonia, or other tone types.

Though only 15/27 subjects had dystonia identified during routine CP Center care using a standardized assessment,^22^ 25/27 had either arm or leg dystonia identified during experts’ consensus-based video review (median GDRS of 1 or more). Of the 10 subjects who did not have dystonia identified in clinic but had a median GDRS of 1 or more after experts’ consensus-building discussions, one subject was rated as not having dystonia by 1 of the 7 experts and another subject was rated as not having dystonia by 3 of the 7 experts. The other 8 subjects who were labeled as not having dystonia in clinic were unanimously rated as having dystonia by the experts. Notably, this is consistent with our previous work demonstrating that, compared to expert consensus-based review, dystonia is under-reported during routine clinical care.^28^ Furthermore, 24/27 subjects had dystonia that functionally impacted the hand open-close or walking task (rated by experts as a 1 or more on the functional impact scale, Table 1), with 11/27 having dystonia that made at least one of the tasks hard to do on their own (rated by experts as a 2 or more). When comparing subjects who did vs. did not have dystonia identified during routine clinical care, there was no significant difference in the experts’ consensus-based GDRS scores or functional impact scores (t-test *p*>0.05, Table 3).

**Table 3.** Experts’ consensus based GDRS and functional impact scores compared between those with and without dystonia per in-clinic evaluation. HOC – alternating hand open-close task.

|  | Overall (n=27) | Dystonia assessment per in-clinic evaluation |  |  |
| --- | --- | --- | --- | --- |
|  |  | No (n=12) | Yes (n=15) | T-test *p* |
| Experts' consensus-based GDRS |  |  |  |  |
| Arm | Average: 5.9<br>95% CI: 4.3 - 7.5<br>Range: 0-14 | Average: 5.5<br>95% CI: 2.6-8.4<br>Range: 0-14 | Average: 6.2<br>95% CI: 4.3-8.1<br>Range: 0-13 | 0.66 |
| Leg | Average: 4.3<br>95% CI: 3.2-5.4<br>Range: 0-11 | Average: 3.8<br>95% CI: 1.7-6.0<br>Range: 0-11 | Average: 4.7<br>95% CI: 3.4-5.9<br>Range: 1-9 | 0.45 |
| Total<br>(Arm + Leg) | Average: 10.2<br>95% CI: 7.9-12.5<br>Range: 0-22 | Average: 9.3<br>95% CI: 4.8-13.9<br>Range: 0-22 | Average: 10.9<br>95% CI: 8.3-13.4<br>Range: 6-19 | 0.51 |
| Experts' consensus-based functional impact score (Table 1) |  |  |  |  |
| HOC | Average: 1.2<br>95% CI: 0.9-1.5<br>Range: 0-2 | Average: 0.9<br>95% CI: 0.4-1.4<br>Range: 0-2 | Average: 1.4<br>95% CI: 1.1-1.7<br>Range: 1-2 | 0.07 |
| Gait | Average: 0.8<br>95% CI: 0.6-1.0<br>Range: 0-2 | Average: 0.8<br>95% CI: 0.4-1.1<br>Range: 0-2 | Average: 0.9<br>95% CI: 0.6-1.2<br>Range: 0-2 | 0.60 |
| Total<br>(HOC + Gait) | Average: 2.0<br>95% CI: 1.6-2.4<br>Range: 0-3 | Average: 1.7<br>95% CI: 0.9-2.4<br>Range: 0-3 | Average: 2.3<br>95% CI: 1.9-2.6<br>Range: 1-3 | 0.11 |

When examining correlations between experts consensus-based arm GDRS, leg GDRS, hand open-close functional impact score, and gait functional impact score, the only significant correlations were between arm GDRS and the hand open-close functional impact score (linear regression R^2^=0.48, *p*=0.0005) and between the leg GDRS and the gait functional impact score (linear regression R^2^=0.43, *p*=0.001). For every 1 point increase in the hand open-close functional impact score, there was a 4.0 point increase in the arm GDRS (Figure 1A). For every 1 point increase in the gait functional impact score, there was a 3.3 point increase in the leg GDRS (Figure 1D).

**Figure 1.**
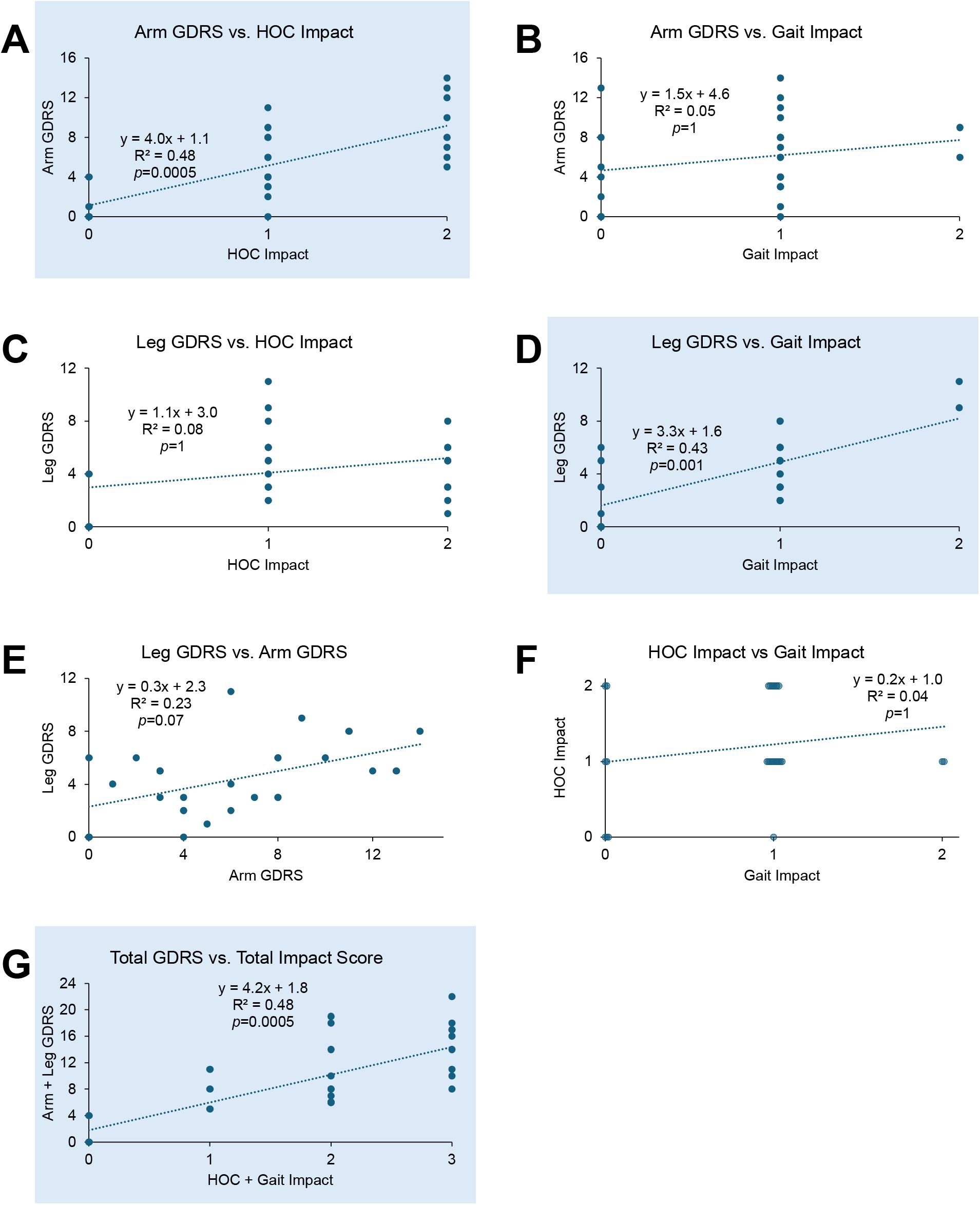
Correlations between experts’ consensus based GDRS and functional impact scores. A-F: Correlations between arm GDRS, leg GDRS, hand open-close (HOC) functional impact score, and gait functional impact scores. G: Correlation between the summed arm and leg GDRS and the summed functional impact score for HOC and gait. (linear regression with Bonferroni corrections for multiple comparisons). Significant correlations (*p*<0.05) are highlighted in blue.

The summed arm and leg GDRS scores also significantly correlated with the summed hand open-close and gait functional impact scores (linear regression R^2^=0.48, *p*=0.0005). For every 1 point increase in the total functional impact score, there was a 4.2 point increase in the total GDRS (Figure 1G).

## Discussion

In ambulatory children with CP, experts’ consensus-based dystonia severity ratings correlated with their assessments of how dystonia functionally impacted two different motor tasks. Arm dystonia severity exclusively correlated with functional impact on an upper extremity task while leg dystonia severity exclusively correlated with functional impact scores on a gait task. Overall, an increase in 4 points on the GDRS correlated with a 1 point increase in functional impact score. This GDRS score difference could be useful for estimating the functionally meaningful difference in dystonia severity in ambulatory children with CP.

Compared to the often-recognized gold-standard (experts’ consensus-based assessment),^25,28,29^ in-clinic evaluation even with a standardized assessment^22^ underestimates the presence of dystonia in children with CP. This replicates our previous work demonstrating clinical under-reporting of dystonia.^28^ Furthermore, the in-clinic evaluation does not just underestimate dystonia in subjects who are mildly impacted, but also in those with more severe dystonia ratings and functional impact. Therefore, dystonia that is not reported or detected during routine clinical care may still be functionally limiting. It thus remains important to use systematic, objective, and even automated strategies to help identify and assess dystonia in people with CP.^25^

Limitations to this study include that results may be generalizable only to ambulatory children with CP. However, these results demonstrate that ambulatory children with CP can still have dystonia that is functionally limiting and may warrant intervention. Future work should examine children with CP who do not independently ambulate. Another limitation is that subject videos were obtained from a single center. However, the 7 expert assessors were from 7 different centers distinct from the center where the videos were obtained. Recruiting experts who cannot have any familiarity with the video subjects ensures unbiased and diverse perspectives on a standardized set of videos (as opposed to experts rating videos of subjects recruited from their own centers). Future work could use both videos and experts from multiple distinct centers while ensuring that experts are not from the same sites generating videos. Finally, as no clinician-assessed functional impact scale for dystonia exists, we had to adapt a caregiver-facing scale for clinicians to directly rate dystonia’s specific functional impact. Future work should cross-validate clinician and caregiver assessments of dystonia’s specific functional impact in the same child.

### Conclusions

We have demonstrated that expert-assessed dystonia severity in children with CP correlates with expert-assessed functional impact and have provided a potential score difference for limb dystonia severity assessment on the GDRS that is associated with a change in functional impact. This score difference could be used as an MCID for the GDRS for ambulatory children with CP. Therefore, this study moves us closer to ensuring that we are assessing functionally meaningful changes in dystonia both in the clinic and in clinical trials.

## Data Availability

All data produced in the present study are available upon reasonable request to the authors

## Notes

### Competing Interest Statement

The authors have declared no competing interest.

### Funding Statement

This study was funded by the Pediatric Epilepsy Research Foundation Registry Planning Grant

